# CHARMS and PROBAST+AI: an updated template for Data Extraction and Risk of Bias Assessment in systematic reviews of prediction models

**DOI:** 10.64898/2026.08.26.26361189

**Authors:** A. Jaber, L.T. Hughes, C. Grimes, A. Vayani, A.C. Cameron, T.J. Quinn

## Abstract

Systematic reviews of clinical prediction models increasingly include studies using artificial intelligence (AI) and machine learning (ML) methods alongside traditional multivariable regression approaches. A previously published Excel tool enabled standardised data extraction using the CHARMS checklist and risk of bias assessment using PROBAST. The recent publication of the PROBAST+AI framework, which distinguishes the assessment of model development quality from the assessment of model evaluation risk of bias and assesses applicability in both parts, necessitates an updated digital instrument applicable across prediction modelling methods.

We updated an open-access Excel tool to incorporate the full PROBAST+AI framework. The updated template incorporates structural separation between assessment of model development quality and model evaluation risk of bias, with applicability assessed in both parts. It also incorporates updated signalling questions, including those addressing methodological issues particularly relevant to AI/ML, and automates the generation of summary tables and graphical displays.

The updated tool (CHARMS & PROBAST+AI Template) contains 12 worksheets and supports data extraction and appraisal for up to 30 prediction models. Dedicated, linked worksheets enable separate assessment of model development and model evaluation, with Domain 4 distinguishing among Apparent, Internal, and External evaluation settings. Key updates include dedicated assessments for predictor pre-processing, class imbalance handling and recalibration, data leakage prevention, and replication of the full model development pipeline within resampling procedures. Automated sheets dynamically format tables and summary charts covering PROBAST+AI parts.

The CHARMS & PROBAST+AI Excel template provides a standardised, user-friendly, and rigorous digital framework for systematic reviewers appraising traditional statistical and AI-driven clinical prediction models.

## Background

Systematic reviews and meta-analyses of clinical prediction models are essential for synthesising evidence on development, evaluation, performance, and applicability of diagnostic and prognostic tools. In 2023, Fernandez-Felix et al. published an open-access Excel template **(1)** to streamline data extraction using the Critical Appraisal and Data Extraction for Systematic Reviews of Prediction Modelling Studies (CHARMS) checklist **(2)** and risk of bias and applicability assessment using the Prediction Model Risk of Bias Assessment Tool (PROBAST)(3). This original tool allowed reviewers to systematically extract item-level study characteristics, evaluate risk of bias across four domains (Participants, Predictors, Outcome, Analysis), assess applicability across the first three domains, and automatically format synthesis tables and summary charts.

Since the publication of the original template, prediction modelling methodology has continued to evolve, alongside the rapid adoption of artificial intelligence (AI) and machine learning (ML) algorithms in clinical medicine (ranging from deep neural networks and gradient-boosted trees to complex high-dimensional feature selection). These developments have highlighted critical gaps that were not explicitly addressed in the original PROBAST framework. Such issues, which are not necessarily exclusive to AI/ML models, include data leakage (4) during feature engineering or splitting, overfitting in high-dimensional space, and distortion of probability calibration caused by synthetic class imbalance corrections, e.g., Synthetic Minority Oversampling Technique (SMOTE) or oversampling (5).

To address these challenges, the PROBAST steering group developed PROBAST+AI, which extends and replaces PROBAST and is applicable to prediction models developed using either regression or artificial intelligence methods(6, 7). In light of this major methodological advancement, we have comprehensively updated the existing Excel template to create the CHARMS & PROBAST+AI extraction template. This article describes the updated structure and signalling questions, dynamic data linking, and automated synthesis capabilities embedded within the updated tool.

## Methodology

### Development of the Updated Template

The updated template retains the user-friendly design and macro-free automation of the original spreadsheet while introducing updates to mirror the PROBAST+AI guidelines. The workbook functions were implemented using native Excel formulas, data validation, conditional formatting, defined names, and protected worksheets, without using Visual Basic for Applications macros. Some updates were made with the assistance of Codex.(8)

### Structural Bifurcation: Development Quality vs. Evaluation Risk of Bias

A fundamental conceptual change in PROBAST+AI is the strict architectural separation between assessing model development and model evaluation (validation). In the original tool, development and evaluation signalling questions were merged within a unified framework. The updated template introduces two distinct worksheets:

- **PROBAST+AI Dev (Development) - Model Development Assessment:** assesses concerns regarding the quality of model development across four domains: Participants and data sources, Predictors, Outcome, and Analysis. Applicability concerns are assessed for the first three domains.
- **PROBAST+AI Eval (Evaluation) - Model Evaluation Assessment:** evaluates the risk of bias and applicability of model evaluation studies. This sheet supports independent assessment across different evaluation settings, including Apparent performance, Internal validation (e.g., cross-validation or bootstrap), and External validation in independent cohorts.

For each included model, users specify whether the publication reports model development only, model evaluation only, or both development and evaluation. This selection determines which assessment worksheet is applicable and how the corresponding results are presented in the automated summaries.

### Multi-Component Evaluation Architecture

The PROBAST+AI Eval worksheet allows reviewers to designate the specific evaluation component for each model assessment. Available options include apparent evaluation only, internal evaluation only, external evaluation only, pairwise combinations of these components, or multiple evaluation components. This structure enables reviewers to assess the methodological limitations of each reported evaluation component while distinguishing performance estimated using the development data from performance estimated through internal resampling or in independent external data.

### Incorporation of new PROBAST+AI Signalling Questions

We incorporated all newly formulated PROBAST+AI signalling questions into Domain 2 (Predictors) and Domain 4 (Analysis). Compared with PROBAST-2019, PROBAST+AI introduces six new signalling questions within the predictors and analysis domains. Although several of these considerations are particularly relevant to complex AI/ML pipelines, their applicability depends on the methods used rather than on whether a model is labelled as statistical or AI-based. These are summarised in Table 1 below.

**Table 1.** New PROBAST+AI signalling questions incorporated into the updated Excel template.

| **Domain** | **PROBAST+AI Item** | **Rationale** |
|---|---|---|
| **Domain 2: Predictors (Dev + Eval)** | 2.2 Predictor Pre-processing<br>*“Was any pre-processing of predictors similar for all participants?”* | Evaluates consistency in pre-processing workflows (e.g., feature scaling, normalization, imputation, or image cropping) across training and evaluation sets. |
| **Domain 4: Analysis (Dev)** | 4.4 Class Imbalance Recalibration<br>*“If methods to address class imbalance were used, was the model or predictions recalibrated?”* | Checks whether artificial balancing (e.g., SMOTE or oversampling) was followed by recalibration to restore baseline prevalence and accurate absolute probability predictions. |
| **Domain 4: Analysis (Eval)** | 4.1 Avoidance of apparent-only evaluation<br>*“Was model evaluation based on only apparent performance avoided?”* | Determines whether model performance was assessed using an appropriate internal or external evaluation method rather than relying exclusively on performance estimated from the model development data. |
| **Domain 4: Analysis (Eval)** | 4.4 Evaluation Class Imbalance<br>*“If methods to address class imbalance were used, was evaluation done without imbalance correction?”* | Ensures model performance metrics (AUC, calibration, Brier score) are evaluated on representative real-world disease prevalence. |
| **Domain 4: Analysis (Eval)** | 4.5 Data Leakage Prevention<br>*“If data splitting was done, was there evidence that data leakage was avoided?”* | Verifies that test set information (e.g., global scaling parameters or repeated measures from the same participant) did not leak into training sets. |
| **Domain 4: Analysis (Eval)** | 4.6 Resampling Pipeline Replication<br>*“If resampling methods were used, were all development steps replicated in resampling?”* | Prevents optimistic performance estimates by verifying that feature selection, hyperparameter tuning, and pre-processing were strictly executed within resampling folds. |

### Independent Validation

The tool was validated by two independent authors from different institutions (CG, AV). CG was recruited from the NHS Fellowship in Clinical AI alumnus network (9). AV was recruited from the Complex Reviews Synthesis Unit (10). Both reviewers were asked to extract data using the tool for an ongoing systematic review(11). They were asked to provide free-form and structured feedback, including cross-checking auto-calculated fields in the tool.

## Implementation

### Workbook Structure

The updated Excel file (named CHARMS & PROBAST+AI Template.xlsx) consists of 12 worksheets (11 visible worksheets and one hidden state-helper worksheet). The introductory worksheet, HOME, provides a general overview of the workbook architecture, usage instructions, and direct links to the relevant CHARMS checklist, PROBAST, and PROBAST+AI methodological papers. (Figure 1)

**Figure 1.**
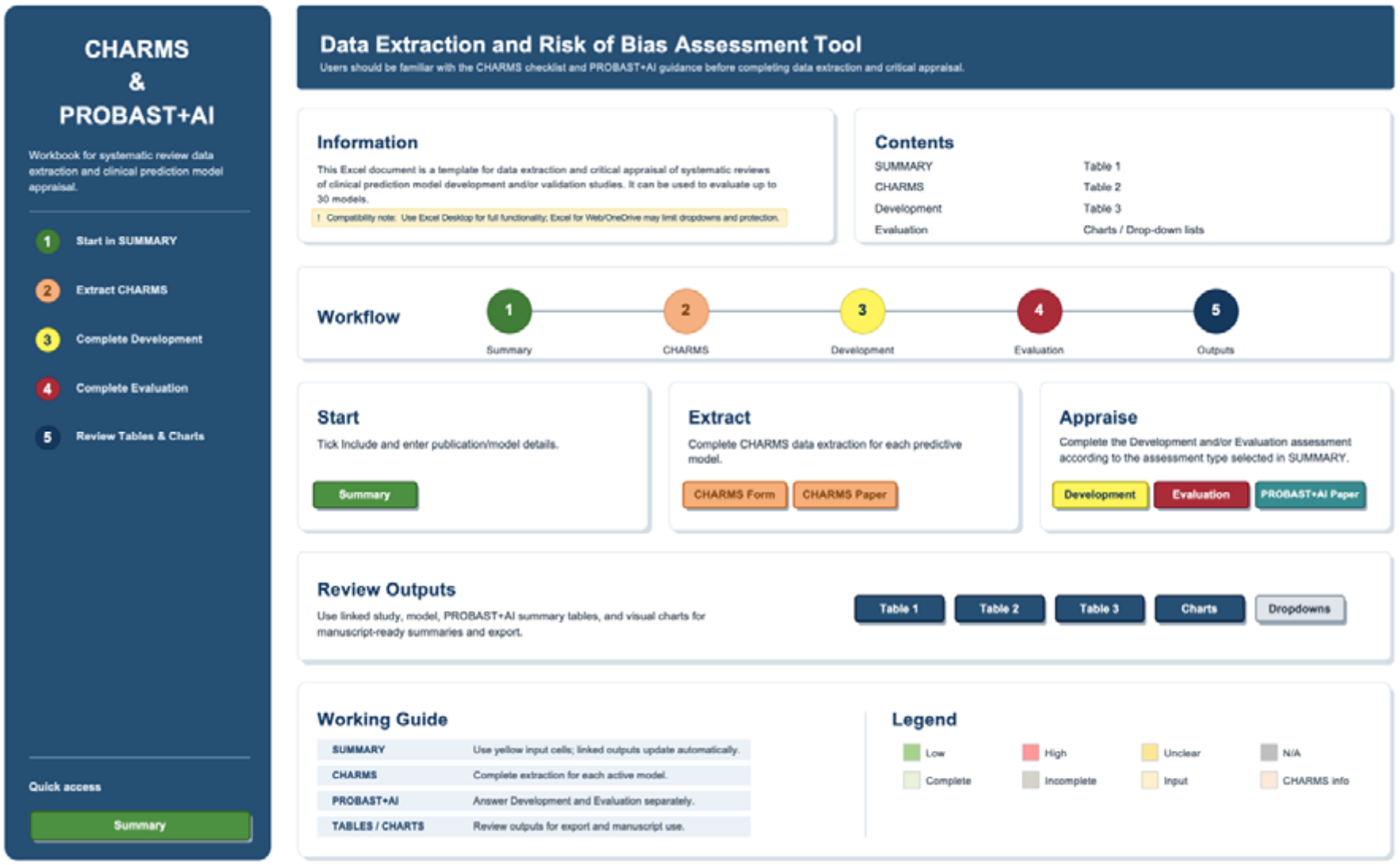
HOME worksheet and overall workflow of the CHARMS & PROBAST+AI Template

The remaining worksheets are divided into three functional layers:

1. **Data Collection & Entry Worksheets:** SUMMARY, CHARMS, PROBAST+AI Dev, and PROBAST+AI Eval.
2. **Automated Synthesis & Reporting Worksheets:** Study Characteristics, Model characteristics, PROBAST+AI summary, and PROBAST+AI Charts.
3. **Template Customisation & Utility Worksheets:** Drop-down lists, version history, and the hidden central helper sheet _MODEL_STATE.

The _MODEL_STATE worksheet manages model indexing, inclusion and exclusion status, display order, assessment type, and the corresponding locations of each model across the extraction and assessment worksheets. These functions are implemented through Excel formulas and defined names and do not require VBA macros.

### Model Activation and Summary Management

The SUMMARY worksheet serves as the central model-management interface. To initiate data extraction for a clinical prediction model presented in an included study, the user selects the tick icon (⍰) from the drop-down menu in the inclusion column on the SUMMARY worksheet. This action activates the corresponding model-specific fields across the extraction and appraisal sheets. The template accommodates up to 30 prediction models per workbook. If a single primary publication reports multiple models (e.g., a baseline regression model and a machine learning model, or distinct development and validation models), the reviewer activates as many model slots as required.

In the SUMMARY sheet, the reviewer completes basic study metadata:

- **Author** (e.g., Ding et al.)
- **Publication Year** (e.g., *2025*)
- **Publication Identifier** (e.g., Title, PMID, or DOI)
- **Publication Journal** (e.g., *BMJ Open Heart*)
- **Model Name** (if applicable)
- **Assessment Type** (e.g., development only)

A composite model identifier (e.g., Ding et al., 2025 – Model Name) is generated automatically from the author, model name and year fields to index the model (or multiple models if reported in the same study or different studies from the same author) across all sheets. The rightmost columns of the SUMMARY sheet provide real-time tracking for each activated model. It displays whether the CHARMS, PROBAST+AI Development, and PROBAST+AI Evaluation sections are *Complete, NA (Not Applicable), or Incomplete*, together with the number of missing required entries in the SUMMARY, CHARMS, and applicable PROBAST+AI assessment fields. It also displays a *Check data* status, which identifies entries that violate predefined validation or consistency rules, such e.g. event count exceeding the corresponding final-analysis sample size. (Figure 2)

**Figure 2.**
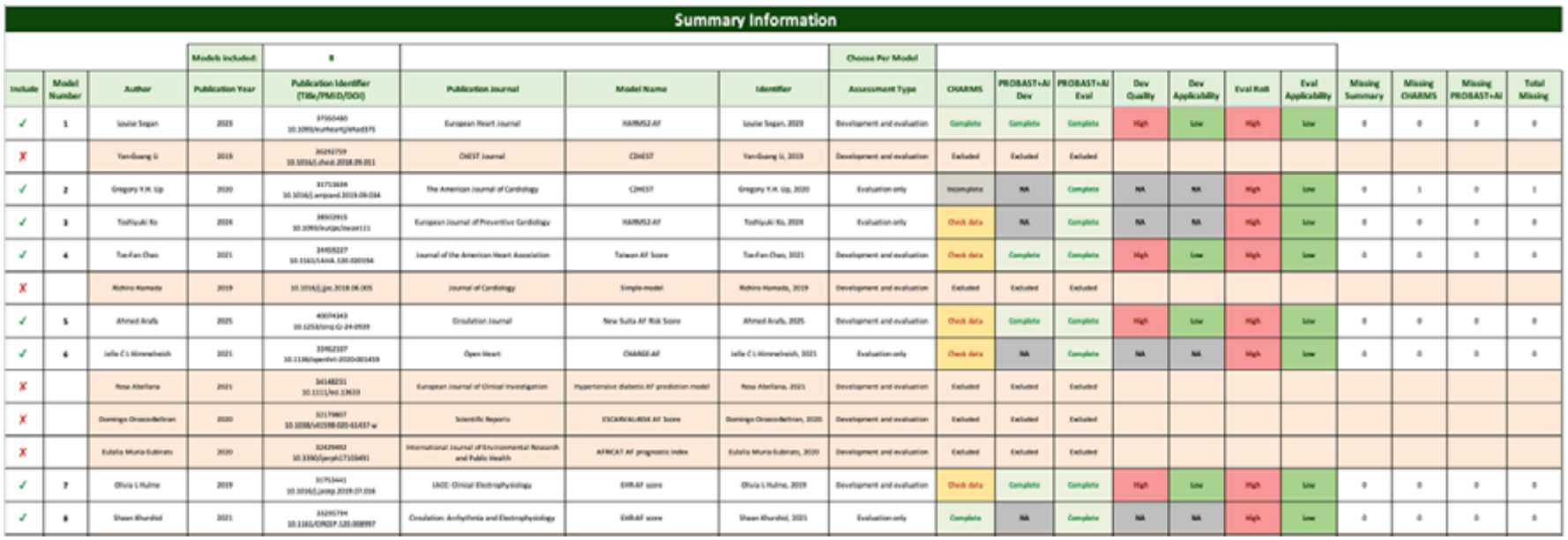
Model management and completion monitoring in the SUMMARY worksheet

Alternatively, if the user decides to exclude a model during or after data extraction and risk of bias assessment, they can select the (X) icon from the drop-down menu in the inclusion column on the SUMMARY worksheet, after which the rightmost columns display the *Excluded* status. The corresponding fields for this model will automatically turn light orange, while the previously entered information remains stored in the workbook. Excluded models are omitted from the automated tables, charts, and included-model counts. Model numbering is recalculated according to the order of the remaining included models, preventing gaps in the output tables.

### Data Extraction

The CHARMS worksheet incorporates the data extraction checklist adapted from Moons et al. (2), updated to capture key machine learning and pre-processing variables. The sheet is organised across 12 extraction domains:

1. *Source of Data*
2. *Participants*
3. *Outcome to be Predicted*
4. *Candidate Predictors*
5. *Sample Size*
6. *Missing Data Handling*
7. *Model Development*
8. *Model Performance*
9. *Model Evaluation*
10. *Results*
11. *Interpretation*
12. *Observations* / *Additional Information*

Reviewers complete all yellow-shaded input cells using standardised drop-down menus or free-text entries. When specific item information is omitted in the primary report, the reviewer selects No information, where available, distinguishing unreported information from fields that have not yet been completed.

Several fields populate automatically based on other fields. These include questions numbered 5.3, 8.1, 8.2, 8.3, and 8.4. Some questions accept numbers or pre-defined text options, such as question 4.1, which accepts whole numbers or ‘Not Applicable’, ‘Unknown’, or ‘No information’. Some questions have a pre-defined drop-down list but also accept free text if the option is not listed. Input messages identify the permitted response format where clarification is useful.

Within the participant-description section, users can define up to five population characteristics according to the review question and apply these consistently across the included models. A domain-level completeness indicator identifies extraction sections with unanswered required fields or illogical answers, and the domain remains marked as *Incomplete* if any mandatory input cell within that domain is left blank or any input needs to be checked. A master status indicator at the base of the CHARMS worksheet flags whether all data fields for a model are fully registered (All required information is complete), pending (Incomplete, with the number of required fields), or need to be checked (Check data, with the number and location of the issues). The number of missing required CHARMS entries is also displayed in the SUMMARY worksheet. (Figure 3) For sections 5 (Sample size) and 6 (Missing data), it is essential that fields are completed in line with the CHARMS guidance **(2)** to ensure that automatic calculations output the correct values.

**Figure 3.**
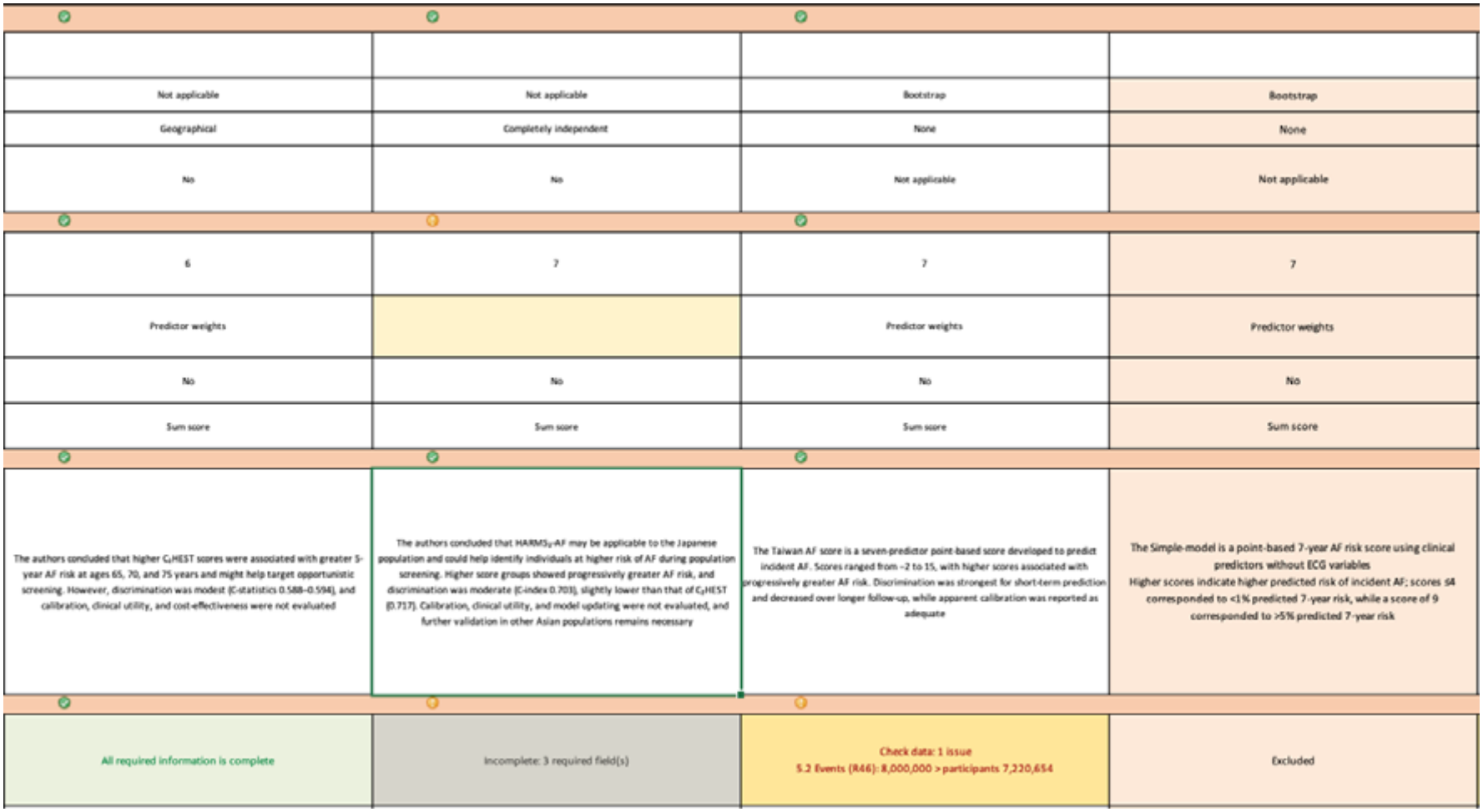
Data extraction and completion monitoring in the CHARMS worksheet

#### Section 5

- 5.1.1 Eligible participants: participants who met the study eligibility criteria before exclusions related to missing data.
- 5.1.2 Participants in the final analysis: participants included in the final model analysis after?exclusions, including exclusions due to missing data.
- 5.2 Number of outcomes/events in the final analysis: the number of outcomes/events among participants included in the final analysis, i.e., among participants counted in 5.1.2.
- 5.3 Number of events per variable/parameter, EPV/EPP: calculated automatically as: (5.2 ÷ 4.1), where 5.2 is the number of outcomes/events in the final analysis and 4.1 is the number of candidate predictors or parameters assessed.

#### Section 6

- 6.1 Handling missing data: the method used to handle missing data. If Complete-case analysis is selected, item 6.2 is activated. If any other method is selected, item 6.3 is activated. If ‘No information’ or ‘Unclear’ are selected, both 6.2 and 6.3 are optional, allowing any available information to be recorded. Selecting ‘Not applicable’ makes neither of them required.
- 6.2 Participants with any missing data among eligible participants: the number of eligible participants with any missing data who were excluded from the final analysis under complete-case analysis.
- 6.3 Participants with any missing data in the final analysis: the number of participantsincluded in the final analysis who had any missing data and were handled using methods other than complete-case analysis, such as imputation or other approaches.

### Model Development Quality Assessment

The PROBAST+AI Development worksheet is applicable to models classified as development only or development and evaluation. It incorporates the 16 PROBAST+AI model development signalling questions across four domains: participants and data sources, predictors, outcome, and analysis. The first three domains contain three, four, and four signalling questions, respectively, while the analysis domain contains five.

To streamline appraisal and eliminate redundant data lookup, relevant extraction items recorded in CHARMS are automatically mirrored into soft rose-shaded reference blocks located directly adjacent to the yellow assessment cells. Reviewers can record additional rationale or notes for each domain where information beyond the linked CHARMS fields is required.

Reviewers will complete yellow-shaded signalling questions across the four core development domains:

- **Domain 1: Participants and Data Sources** (Items 1.1–1.3)
- **Domain 2: Predictors** (Items 2.1–2.4, including Item 2.2 on predictor pre-processing consistency)
- **Domain 3: Outcome** (Items 3.1–3.4)
- **Domain 4: Analysis** (Items 4.1–4.5, including Item 4.4 on probability recalibration following class imbalance correction)

Responses are selected from drop-down menus containing Y (*Yes*), PY (*Probably yes*), PN (*Probably no*), N (*No*), or NI (*No information*). NA (*Not applicable*) is available for questions explicitly identified by PROBAST+AI as conditional. Based on these signalling questions, reviewers assign overall domain-level ratings for **Quality Concerns** and **Applicability Concerns** as Low (+), High (-), or Unclear (?). (Figure 4)

**Figure 4.**
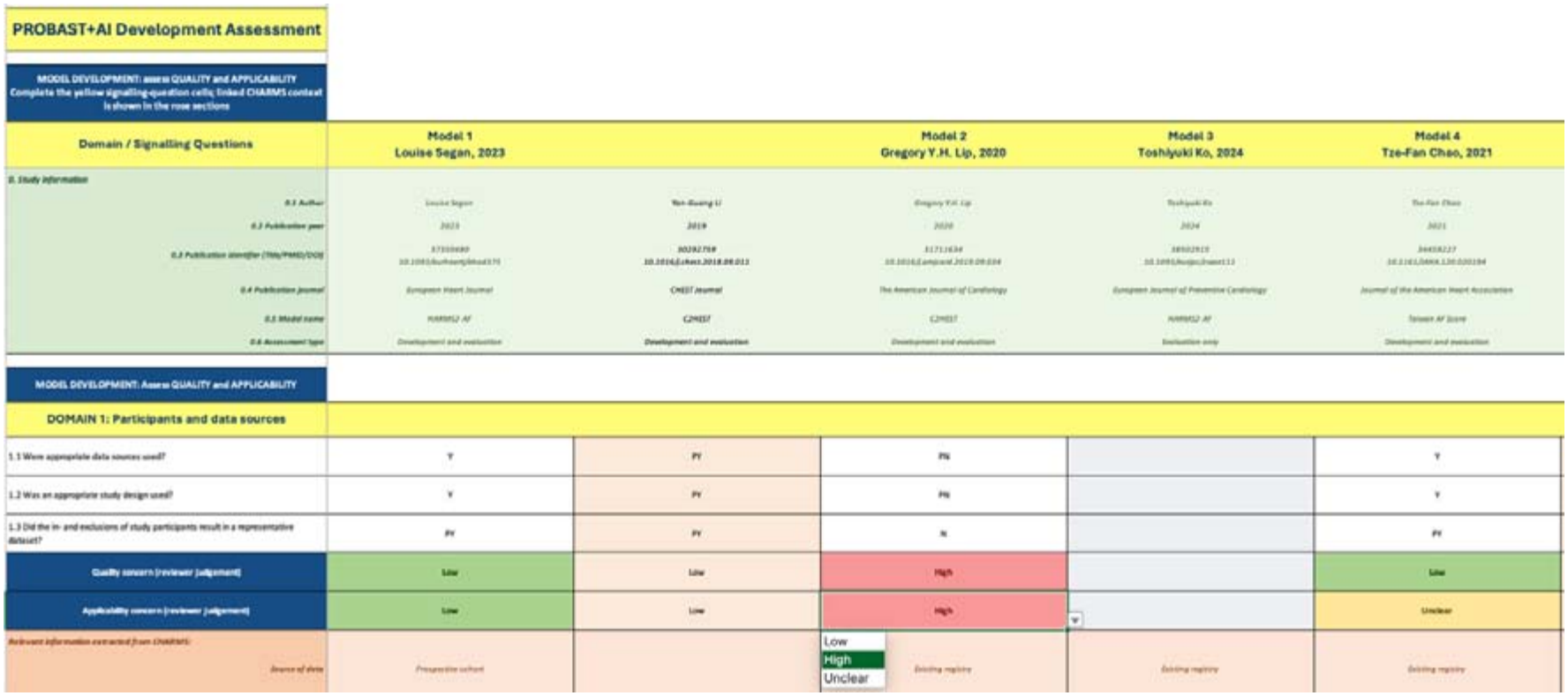
Model development quality and applicability assessment in the PROBAST+AI Development worksheet

The overall development quality and applicability judgements are generated automatically from the reviewer-assigned domain judgements. An overall judgement is rated high if at least one relevant domain is rated high, unclear if at least one domain is rated unclear and none is rated high, and low when all relevant domains are rated low. For models classified as evaluation only, the development assessment is reported as NA (Not applicable).

### Model Evaluation Risk of Bias Assessment

The PROBAST+AI Evaluation worksheet is applicable to models classified as evaluation only or development and evaluation. It incorporates the 18 PROBAST+AI model evaluation signalling questions across four domains: participants and data sources, predictors, outcome, and analysis. The first three domains contain three, four, and four signalling questions, respectively, while the analysis domain contains seven.

As in the development sheet, extracted CHARMS data are displayed in rose-coloured context panels. Reviewers answer the signalling questions using Y (*Yes*), PY (*Probably yes*), PN (*Probably no*), N (*No*), NI (*No information*), or NA (*Not applicable*) where permitted. Reviewers then record domain-level **Risk of Bias** and **Applicability** ratings as Low, High, or Unclear.

The PROBAST+AI Eval worksheet assesses risk of bias (RoB) and applicability concerns for model evaluation or validation components. Crucially, this sheet supports multi-setting evaluation by allowing reviewers to specify the evaluation type assessed in Item 4.0:

- **Apparent Only** (performance on development data)
- **Internal Only** (e.g., cross-validation, bootstrap)
- **External Only** (independent validation cohort)
- **Pairwise combination from the above three**
- **Multiple** (all three types of validation)

The selected classification determines which apparent, internal, and external signalling-question fields are applicable and which fields are included in the automated completeness assessment. Questions that are not relevant to a selected evaluation component are treated as not applicable. Reviewers complete signalling questions across the four evaluation domains:

- **Domain 1: Participants and Data Sources** (Items 1.1–1.3)
- **Domain 2: Predictors** (Items 2.1–2.4)
- **Domain 3: Outcome** (Items 3.1–3.4)
- **Domain 4: Analysis - Model Evaluation** (Items 4.1–4.7, including Item 4.4 on unadjusted evaluation datasets, Item 4.5 on data leakage prevention, and Item 4.6 on full pipeline replication within resampling)

Within each model record, Domains 1-3 and item 4.1 are shared across the selected evaluation components. Applicable questions in items 4.2-4.7 provide separate response fields for apparent, internal, and external evaluation. Reviewers assign one risk-of-bias judgement for Domain 4, and the workbook generates one overall evaluation risk-of-bias judgement and one overall applicability judgement per model record.

Overall evaluation risk of bias and applicability are generated automatically from the reviewer-assigned domain judgements. Overall risk of bias is rated high when at least one domain is rated high, unclear when at least one domain is rated unclear and none is rated high, and low when all four domains are rated low. The same rule is applied to the three applicability domains. For models classified as development only, the evaluation assessment is reported as NA (Not applicable).

When all development quality and evaluation RoB domains are scored, the model’s overall assessment status is updated automatically in the SUMMARY sheet.

## Results

### Automated Reporting, Visualisation, and Customisation

The final seven worksheets format the data collected only from the activated models into outputs and manage workbook customisation:

- Study Characteristics **(Table 1):** Automatically aggregates primary study design, enrolment dates, clinical setting, geographic region, and participant demographic summaries across included models. When multiple prediction models were reported in the same publication, information was presented at the model level and could therefore appear in more than one output row. (Figure 5)
- Model characteristics (**Table 2**): Compiles algorithm types (e.g., Logistic Regression, Neural Networks, Random Forests), sample sizes, event counts, candidate vs. final predictor counts, missing data strategies, validation types, and performance metrics. When numeric data are available, the table automatically calculates event percentages using the number of participants in the final analysis as the denominator. Missing-data percentages are calculated according to the reported missing-data approach: for complete-case analysis, the denominator is the number of eligible participants; for other missing-data methods, the denominator is the number of participants in the final analysis. EPV/EPP is calculated automatically as the number of events divided by the number of candidate predictors or parameters assessed. (Figure 6)
- PROBAST+AI summary (**Table 3**): Generates a complete domain-by-domain matrix summarising **Development Quality, Development Applicability, Evaluation Risk of Bias**, and **Evaluation Applicability** for every included model. (Figure 7)
- PROBAST+AI Charts: Generates stacked bar charts and traffic-light summary figures summarising development quality, development applicability, evaluation risk of bias, and evaluation applicability. For each domain or overall assessment, percentages are calculated from included records with an applicable Low, High, or Unclear judgement. Excluded records, non-applicable assessments, and missing judgements are omitted from the corresponding denominator. The number of contributing records may therefore differ between charts. (Figure 8)
- Drop-down lists: Contains the controlled vocabularies for CHARMS and PROBAST+AI drop-down menus.
- Version history: Identifies the workbook version and summarises changes from the previous version.
- _MODEL_STATE: A hidden system sheet that manages internal model indexing, visibility order, and logical state verification without requiring VBA macros.

**Figure 5.**
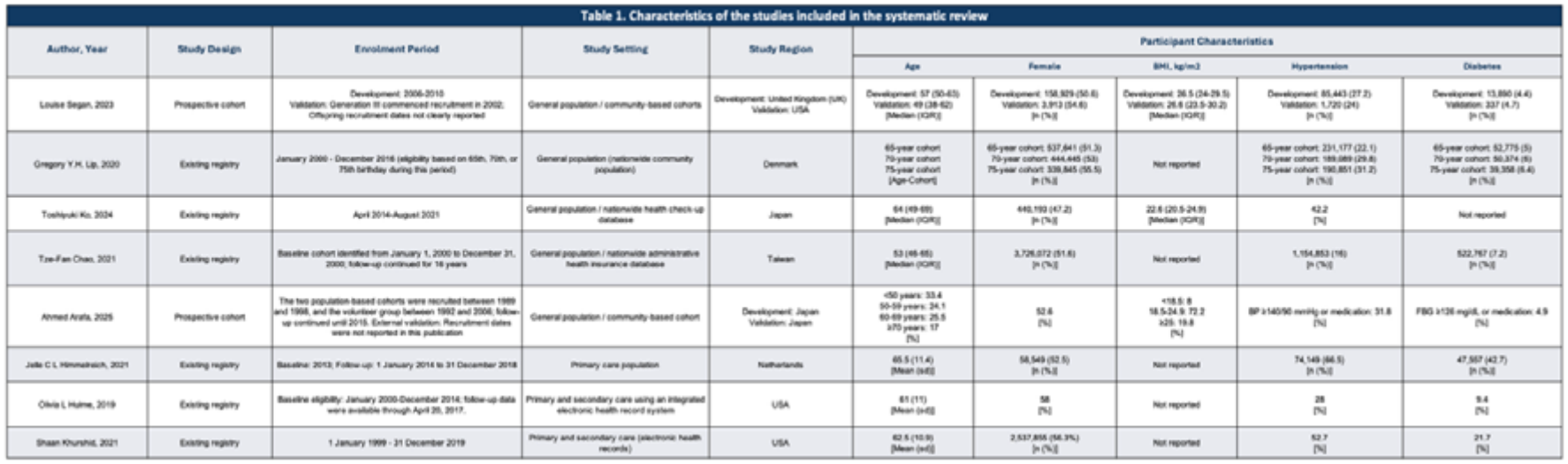
Automatically generated Study Characteristics table from the illustrative dataset

**Figure 6.**
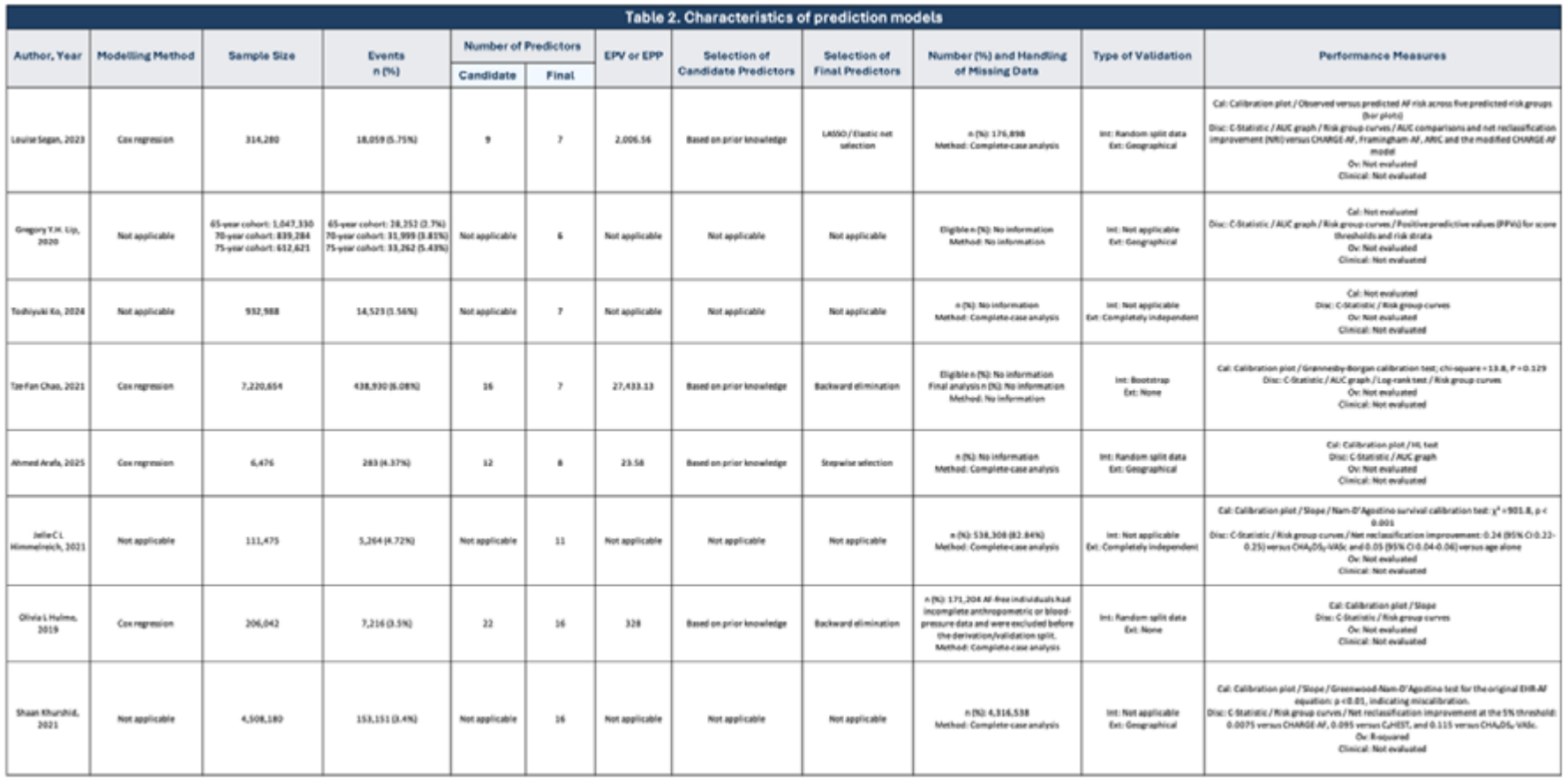
Automatically generated Model Characteristics table from the illustrative dataset

**Figure 7.**
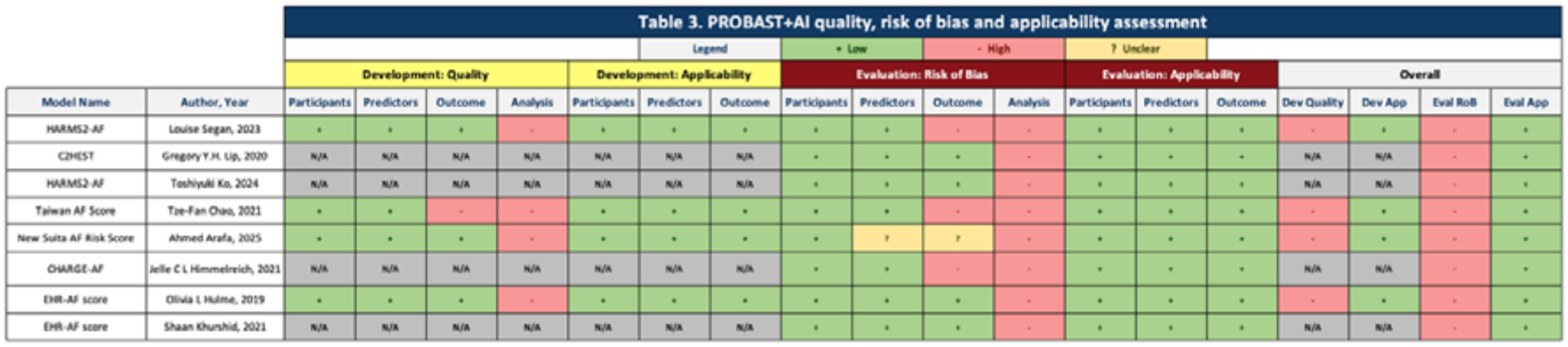
Automatically generated PROBAST+AI summary table from the illustrative dataset

**Figure 8.**
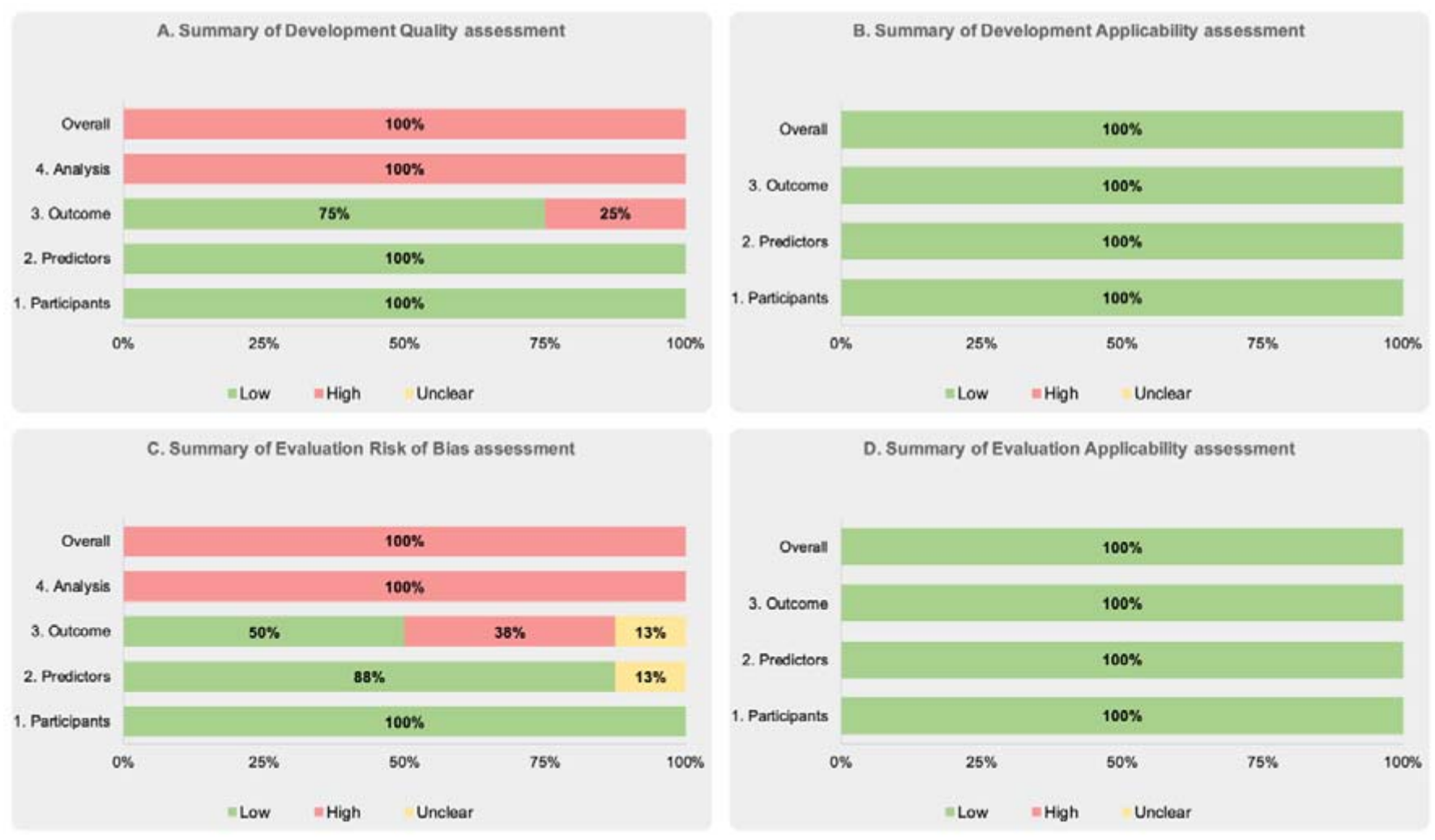
Automatically generated PROBAST+AI graphical summaries from the illustrative dataset

### Dynamic Printing and PDF Export

The Study Characteristics, Model Characteristics, and PROBAST+AI Summary worksheets use dynamic print areas. These print areas automatically adjust according to the number of models included in the synthesis, ensuring that populated rows are included while unused rows are omitted from the printed output.

The automatically generated tables are protected against editing, although users can select and copy their contents. Users can preview the dynamically formatted outputs by opening the system print dialogue using Command+P on macOS or Ctrl+P on Windows. The resulting tables can then be printed or exported as PDF files using the available system print options. This allows the automatically generated tables to be incorporated into manuscripts, reports, or supplementary materials without manually defining the required print range

### Illustrative Application

To demonstrate the practical application of the updated template, a completed example based on data extracted from an ongoing systematic review of atrial fibrillation prediction models is provided as Supplementary File S2. The example contains 13 completed model records, of which eight were selected for inclusion in the automated outputs. It includes studies reporting model development and evaluation as well as studies reporting evaluation only.

The completed workbook demonstrates how the ‘SUMMARY’ worksheet manages assessment type, model inclusion, and completion status; how study and model information is recorded in the CHARMS worksheet; and how extracted information is linked to the corresponding PROBAST+AI Development and Evaluation assessments. It also demonstrates the automatic generation of study characteristics, model characteristics, PROBAST+AI summary tables, and graphical displays. Models excluded from the synthesis remain stored in the workbook but are omitted from the generated outputs.

### Independent Validation

Feedback from the Independent Validation conducted by CG and AV was reviewed by LH and AJ. Feedback was itemised and categorised by: Tool Functionality Problems; Excel Limitation; User Experience Factor. This led to ‘bugs’ being fixed, additional features being implemented, and guidance notes being included. Certain feedback elements would have improved usability for individual authors but may have reduced the tool’s wider applicability or departed from the frameworks from which it was developed. Decisions to include or not include features were at the discretion of two authors (AJ, LH).

## Discussion

This work provides a revised and organised Excel template that combines CHARMS data extraction with the new PROBAST+AI framework. The main improvement over the original CHARMS and PROBAST template is the clear separation of model development quality from the risk of bias in model evaluation. There are also separate assessments of applicability for both parts. This separation is important because the quality of the model development process and the risk of bias in the evaluation can differ significantly within the same study or model.

The updated template also addresses methodological issues that have become more important with the rise of artificial intelligence and machine learning in clinical prediction modelling. These issues include preparing predictors, correcting class imbalance and recalibrating, avoiding data leakage, and replicating the entire model development process during resampling. However, the template is not limited to AI or machine learning models. It can also be used for prediction models created with traditional multivariable regression methods. This helps reviewers apply a consistent extraction and evaluation framework across systematic reviews that include various modelling techniques.

The workbook aims to support, not replace, methodological judgment. Reviewers are still responsible for interpreting the signalling questions, considering additional information in the source publications, and assigning quality, risk-of-bias, and applicability judgments at the domain level.

By transforming the PROBAST+AI framework into a structured and user-friendly digital format, the updated template may encourage more consistent data extraction and critical evaluation across systematic reviews of prediction models. It could be especially useful for reviews that include both traditional statistical models and AI or machine learning methods, as it offers a common framework to address their shared and unique concerns.

The tool will be hosted within the Complex Reviews Synthesis Unit for ongoing access. Feedback is welcomed and should be directed to the corresponding author.

## Limitations

Despite these significant structural and functional enhancements, several software-based, methodological, and workflow limitations of the updated template should be acknowledged:

- The current template remains pre-configured to accommodate a maximum of 30 prediction models per workbook instance. Managing larger volumes of models may require partitioning across multiple workbook instances. Systematic reviews containing more than 30 models would therefore require the use of additional workbook copies or modification of the existing workbook structure.
- As a standalone Excel workbook, the tool does not natively support dual extraction or automated inter-author conflict resolution. Independent extraction by two reviewers necessitates manual file comparison.
- To maximise accessibility without security alerts, the workbook operates without macros. This relies on nested Excel cell formulas and relative referencing. Accidental row deletion, improper copy-pasting, or overwriting calculated fields can disrupt the dynamic links between the CHARMS extraction sheet and PROBAST+AI assessment panels. Although calculated cells and output worksheets are protected against unintended editing, removing worksheet protection, modifying the workbook structure, or using unsupported copy-and-paste operations may disrupt the links between extraction, assessment, and output worksheets. Some formatting, formula, chart, protection, or dynamic-printing features may also behave differently in spreadsheet software other than Microsoft Excel.
- The tool is designed to be used in the desktop Windows or macOS version of Excel. The tool may not function correctly in web-based spreadsheet tools.

## Conclusion

By providing a standardised, macro-free, open-access spreadsheet that supports both conventional multivariable regression and complex AI architectures, this tool aims to reduce reviewer workload, improve extraction accuracy, and promote transparent risk-of-bias reporting in line with PRISMA guidelines. The separation of development quality and evaluation risk of bias ensures that systematic reviewers can accurately evaluate the methodological rigor of model creation independently from its validation.

User feedback should be directed to the corresponding author.

## Supporting information

CHARMS & PROBAST+AI Template - Blank

CHARMS & PROBAST+AI Template - Pre-filled Example

## Data Availability

All data produced in the present study are included in the manuscript and supplementary material.

## Authors’ contributions

A. Jaber devised and designed the updated template. A. Jaber, L. T. Hughes, C. Grimes and A. Vayani contributed feedback to the tool’s development. C. Grimes and A. Vayani completed independent validation of the template’s function. The original draft was written by L. T. Hughes and A. Jaber. A. Jaber, L. T. Hughes, C. Grimes, A. Vayani, A. C. Cameron, and T.J. Quinn contributed to reviewing and editing the article. All authors read and approved the final manuscript.

## Declarations

Availability of data and materials: The updated Excel spreadsheet (CHARMS & PROBAST+AI Template.xlsx) is freely available for download and open-source research use under Creative Commons License – CC BYSA

### Use of AI

Artificial Intelligence (Gemini) was used to assist with formatting and proof-reading. The authors have read and take full responsibility for the manuscript.

